# ROLE OF cfDNA AND ctDNA TO IMPROVE THE RISK STRATIFICATION AND THE DISEASE FOLLOW-UP IN PATIENTS WITH ENDOMETRIAL CANCER: TOWARDS THE CLINICAL APPLICATION

**DOI:** 10.1101/2024.05.20.24307623

**Authors:** Carlos Casas-Arozamena, Ana Vilar, Juan Cueva, Efigenia Arias, Victoria Sampayo, Eva Diaz, Sara S Oltra, Cristian Pablo Moiola, Silvia Cabrera, Alexandra Cortegoso, Teresa Curiel, Alicia Abalo, Mónica Pamies Serrano, Santiago Domingo del Pozo, Pablo Padilla-Iserte, Marta Arnaez de la Cruz, Alicia Hernández, Virginia García-Pineda, Juan Ruiz Bañobre, Rafael López, Xavier Matias-Guiu, Eva Colás, Antonio Gil-Moreno, Miguel Abal, Gema Moreno-Bueno, Laura Muinelo-Romay

## Abstract

**Introduction:** In the past years, there has been a rise on advanced endometrial cancers (EC) patients resulting in mortality increase. To overcome this trend, it is essential to improve the stratification of the risk of post-surgery recurrence and to anticipate the development of disease relapse and resistance to treatment. Liquid biopsy analyses represent a promising tool to address these clinical challenges, however, the best strategy to efficiently apply them in the context of EC must be better defined. Therefore, the study was designed to determine the value of cfDNA/ctDNA monitoring to improve the clinical management of patients with localized and recurrent disease.

**Material & Methods:** Plasma samples and the uterine aspirate (UA) from 198 patients with EC were collected in different Spanish hospitals at surgery and throughout the course of the disease. The genetic landscape of UAs was characterized using targeted sequencing. Total cfDNA was isolated from all plasma samples, quantified, and analysed for the presence of ctDNA based on the mutational profile found on the UAs.

**Results:** The genetic characterization of UAs obtained at surgery allowed the identification of pathogenetic variants in the 95,45% of the tumours and ctDNA levels could be monitored in the 89,4% of the patients. High levels of cfDNA and detectable levels of ctDNA at baseline correlated with poor prognosis, for both DFS (p-value<0.0001; HR=9,25) and DSS (p-value<0.0001; HR=11,20). Importantly, this approach remains clinically significant when stratifying tumours based on histopathological risk factors, highlighting its additional value to identify patient with a poor evolution. In fact, cfDNA/ctDNA analysis served to identify patients who showed early post-surgery relapse. Moreover, longitudinal analyses of cfDNA/ctDNA proved to be a powerful asset to identify patients undergoing relapse, months prior to the arisen of any clinical evidence.

**Conclusion:** This study represents the most comprehensive study on cfDNA/ctDNA characterization in EC and demonstrates its value to improve the risk stratification and anticipate the disease relapse in patients with localized disease. Besides, the dynamic ctDNA assessment showed utility to complement the current strategies to monitor disease evolution and the response to treatment. Implementation of cfDNA/ctDNA monitoring into the clinical routine will provide an unique opportunity to improve EC management.

**GRAPHICAL ABSTRACT:** 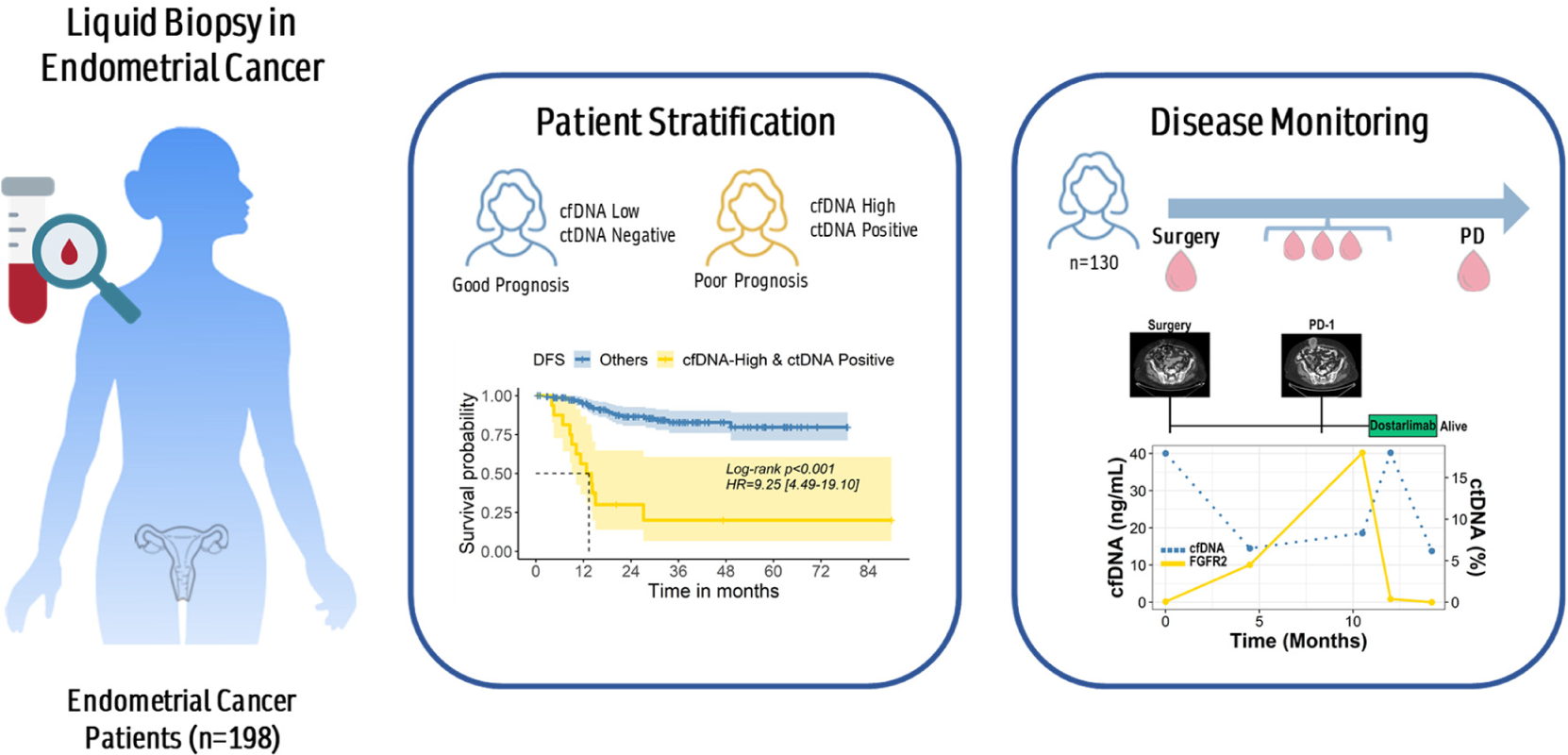

## 1. INTRODUCTION

In the past years, there has been a rise on advanced endometrial cancers (EC) patients resulting in an increase on the EC mortality rates (1,2). Moreover, currently available therapeutic approaches for EC patients have shown limited efficiency on advanced disease (3). About 20% of EC patients will develop recurrent disease or distant metastases (1) after primary treatment, being the identification of patients at higher risk of relapse a clear unmet need. Currently, pathological and molecular information are used to predict prognosis. Characteristics such as high-grade, advanced FIGO stage, non-endometrioid histology and the combination of these features with molecular subgroups has refined the prognostic prediction (1,4–6). In patients with high-intermediate/high risk tumours, clinical guidelines recommend adjuvant treatment to eliminate potential post-surgery residual disease (5). However, only part of these patients really benefit from the adjuvant treatment since over 80% are considered as cured after the surgery (7). Thus, more precise tools are required to improve the risk stratification and the selection of patients that should receive adjuvant therapy.

Another key challenge is the sequential therapy. Although EC treatment is evolving and the immunotherapy and targeted therapies are gaining more interest, the selection of the best treatment sequence/combination for each patient at the best moment requires precise follow-up tools to anticipate the emergence of resistance (1,8). Tumour heterogeneity and clonal evolution in response to therapy are also key milestones to overcome in order to improve EC patient management (9,10). In fact, the molecular characterization of uterine aspirates (UAs) from EC patients has clearly showed the relevance of minimally invasive samples to capture the genetic heterogeneity present in the primary tumour (9,10). Besides, the analysis of circulating biomarkers such as circulating cell free DNA (cfDNA), allows the dynamic characterization of the tumour with minimal discomfort for the patients, and provide with valuable real time information about the disease evolution (11–16). Although few studies have been published focusing on the value of liquid biopsy-based approaches in EC, they showed promising results (17,18). For instance, higher levels of cfDNA have been associated to high risk tumours (19–25). Moreover, Bolivar et al have shown that high levels of cfDNA correlate with worse disease free survival (DFS) and disease specific survival (DSS) (23). Regarding the specific tumour derived cfDNA (ctDNA), our group reported around 40% of patients with EC with detectable levels of ctDNA at surgery, being the patients with the higher risk of recurrence those with higher levels of ctDNA (19).

Considering all this scientific and clinical context and advance towards the application of liquid biopsy in EC, current study was designed to determine the value of cfDNA/ctDNA to complement the current risk stratification and to assess their capability to anticipate the disease relapse with the final goal to improve the clinical management of patients with localized and recurrent disease.

## 2. MATERIAL AND METHODS

### 2.1 Patients’ inclusion and sample collection

A total of 198 patients with EC were recruited between January 2018 and June 2022 at the Gynecology Department of Vall d’Hebron University Hospital (Barcelona, Spain), the MD Anderson Cancer Center (Madrid, Spain), the University Clinical Hospital of Santiago de Compostela (Santiago de Compostela, Spain) and the University Hospital La Fe (Valencia, Spain).

Patients were included into the study if the following criteria were met: (a) patients diagnosed with endometrial adenocarcinomas of any histology; (b) patients older than 18 years; (c) patient’s signed informed consent; (d) patients were not undergoing antitumoral treatment at the time of sample collection; (e) patients did not have any cancer within the last 5 years before sample collection.

All UAs were collected at surgery using a Cornier canula and processed as previously described (26). Peripheral blood samples were collected using CellSave Preservative tubes (Silicon Biosystems Inc, Huntington Valley, USA). Plasma was isolated by a two-step centrifugation (1500xg and 5500xg, respectively). Longitudinal peripheral blood samples were mainly collected every 6 months for 2 years after surgery, and when a recurrence was suspected or confirmed by imaging and/or biopsy. In a subgroup of 37 patients, blood was also obtained 1 month post-surgery, in addition to the mentioned follow-up scheme.

### 2.2 Nucleic acids isolation

DNA and RNA from the UA were obtained using RecoverAll™ Total Nucleic Acid Isolation Kit (Thermo Fisher Scientific, Waltham, MA, USA) following the manufacturer’s conditions. DNA from plasma samples was extracted with the QIAamp DNA Circulating Nucleic Acid Kit (Qiagen, Venlo, Netherlands), according to the manufacturer’s instructions. DNA and RNA from FFPE samples were isolated using the AllPrep DNA/RNA FFPE Kit (Qiagen, Venlo, Netherlands). All DNA samples were quantified using the Qubit Fluorometer (Thermo Fisher Scientific, Waltham, MA, USA) and stored at −20°C until use.

### 2.3 CfDNA characterization by ddPCR

For each patient, specific ddPCR assays were designed based on the genomic landscape identified in the UA and run on a QX-200 dPCR system (Bio-Rad, California, USA). PCR was performed with the ddPCR Supermix for probes (Bio-Rad, Santa Rosa, CA, USA). The sample was partitioned into a median of 50,000 droplets (run in triplicates) in an automated droplet generator (Bio-Rad, CA, USA), according to the manufacturer’s instructions. Emulsified PCR reactions were run on 96-well plates on a C1000 Touch^TM^ thermal cycler (Bio-Rad, CA, USA) according to the manufacturer’s instructions. Plates were read on a Bio-Rad QX-200 droplet reader with Bio-Rad’s QuantaSoft v1.7.4 software to quantify the number of droplets positive for mutant DNA, wild-type DNA, both, and neither. Analysis was performed manually by two independent molecular biologists according to the following guidelines: a minimum of 30,000 positive droplets across wells were required for a valid assay, and a minimum of five, single FAM-positive or HEX-positive droplets with no positive events in the WT control were required to consider samples as mutated.

### 2.4 Targeted sequencing of the uterine aspirate

The UAs underwent targeted sequencing using the Oncomine Comprehensive Panel v3 (Thermo Fisher, Pleasanton, CA,USA) following previously published protocols (19,26). This panel comprises 161 genes categorized by somatic alteration type, including 87 Hotspots genes, 43 focal CNV gains, 48 Full CDS for DEL mutations, and 51 fusion drivers, which cover all the most frequent mutations found in EC.

In summary, 10 ng of both DNA and cDNA from each UA were utilized for library assembly via multiplex PCR on an AB2720 Thermal Cycle (Life Technologies, Carlsbad, California, USA), adhering to the manufacturer’s instructions. PCR amplification involved 18 and 20 cycles. Subsequently, primary primers underwent partial digestion using FuPa reagent (Thermo Fisher, Pleasanton, CA, USA). Ion P1 Adapter and Ion Xpress Barcode X were employed for amplicon ligation, followed by library purification and quantification using the Ion Library TaqMan Quantitation Kit and ViiA 7 system. Libraries were diluted to match the concentration range of the Escherichia coli DH10B Control Library standards, with relative concentration determined through qPCR analysis. Template preparation and enrichment were conducted using the Ion S5 XL system. Diluted libraries were combined with template-positive Ion Sphere Particles (ISPs) and Ion S5 enzyme mix for emulsion PCR, followed by enrichment on the Ion OneTouch 2. Targeted massive sequencing was performed on the S5 sequencer (Thermo Fisher, Pleasanton, CA, USA) with six libraries (RNA and DNA) run in 540 chips. Duplicates were analysed for 10% of the samples and yielded consistent results.

For the bioinformatics analysis, alignment to the Hg19 human reference genome and variant calling were executed using Torrent Suite Software v.15.1 (Life Technologies, Carlsbad, California, USA). Variants with a Phred quality score field value <100 were considered low-quality, while the prediction of genomic variant effects on protein function was conducted using the PROVEAN Genome Variants tool (http://provean.jcvi.org/index.php) and Alamut Visual Plus. Variants predicted as possibly damaging or deleterious by at least one PROVEAN predictor were visually inspected with Integrative Genomics Viewer (IGV) v.2.3.40, Broad Institute. Variants with a global minor allele frequency above 0.05 were categorized as single nucleotide polymorphisms and excluded (data from dbSNP, [http://www.ncbi.nlm.nih.gov/SNP/]).

### 2.5 Statistical Analysis

Statistical analysis was conducted in R (R Core Team, 2020) and figures were generated using ggplot2 (122) and GraphPad Prism 8.0 (GraphPad Software, Inc., San Diego, CA, USA). A Cox-proportional hazard model was used to determine the correlation of clinical and experimental variables with clinical outcomes. Wilcoxon’s signed-rank test was used to evaluate statistical differences in non-parametric experimental variables. The Spearman correlation test was performed to determine the relationship between experimental nonparametric variables. The Pearson correlation test was performed to determine the relationship between parametric quantitative experimental and clinical variables. Associations between clinicopathologic features and the experimental variables were examined with the chi-square test (Fisher’s exact test). The RegParallel package (28) was used to stablish the optimal cut-point to determine the cfDNA utility as a predictor of poor clinical outcome. A P-value <0.05 was set as the level of statistical significance.

## 3. RESULTS

### 3.1. Clinicopathologic Characteristics of the cohort

A total of 198 patients with EC and with at least 6 months of post-surgery follow up have been prospectively included in the study. Clinical characteristics are summarised in Supplementary Table 1. The cohort included patients with endometrioid (76%) and non-endometrioid carcinomas (24%), low (G1-2) and high grade (G3) (58% and 42%, respectively), FIGO stage I-IV (65%, 15%, 15%, 4.7%, respectively) tumours from all TCGA groups (POLE 7,7%, MSI 39%, NSMP 27%, HCN 26%) classified accordingly with the updated ESMO/ESGO consensus (1). A total of 37 (19%) patients had a relapse with a median DFS of 13.9 months [2.6-49.2]. Twenty-four patients (12%) died of disease, showing the global cohort a median DSS of 19.1 months [8.6-45.7].

### 3.2. Targeted sequencing of UAs for personalized ctDNA detection

The UAs from all the patients were subjected to NGS using a targeted panel previously used to characterize EC patients (10,26). With this strategy we identified pathogenic mutation in 189 of the patients (95.46%) The 10 most commonly mutated genes for SNPs were *PTEN* (54.46%), *PIK3CA* (48.51%), *TP53* (30.69%), *ARID1A* (28.71%), *KRAS* (20.79%), *CTNNB1* (19.31%), *PIK3R1* (17.82%), *FBXW7* (14.36%), *PPP2R1A* (13.37%) and *FGFR2* (9.41%). When focusing on the CNAs, 19 patients (9.59 %) had CNAs. The top 10 altered genes for CNAs are *CCNE1* (4.46%), *ERBB2* (2.48%), *CDK2* (1.98%), *AKT2* (0.99%), *MDM2* (0.99%), *MYC* (0.99%), *PIK3CA* (0.99%), *AR* (0.50%), *AXL* (0.50%) and *CCND3* (0.50%). These data are in line with the most frequent alterations of EC described in tissue and UAs samples (26,29).

Based on the genetic alterations found in the UAs specific ddPCR were design to monitor those with the highest allelic frequency. In MSI tumours a panel of 5 microsatellite markers was also assessed by ddPCR. Overall, we could design effective assays to monitor ctDNA in 177 patients. In addition to the pre-surgery point, we obtained follow-up blood samples from 130 patients (65.66%) to study the value of longitudinal cfDNA/ctDNA monitoring (Figure 1).

**Figure 1:**
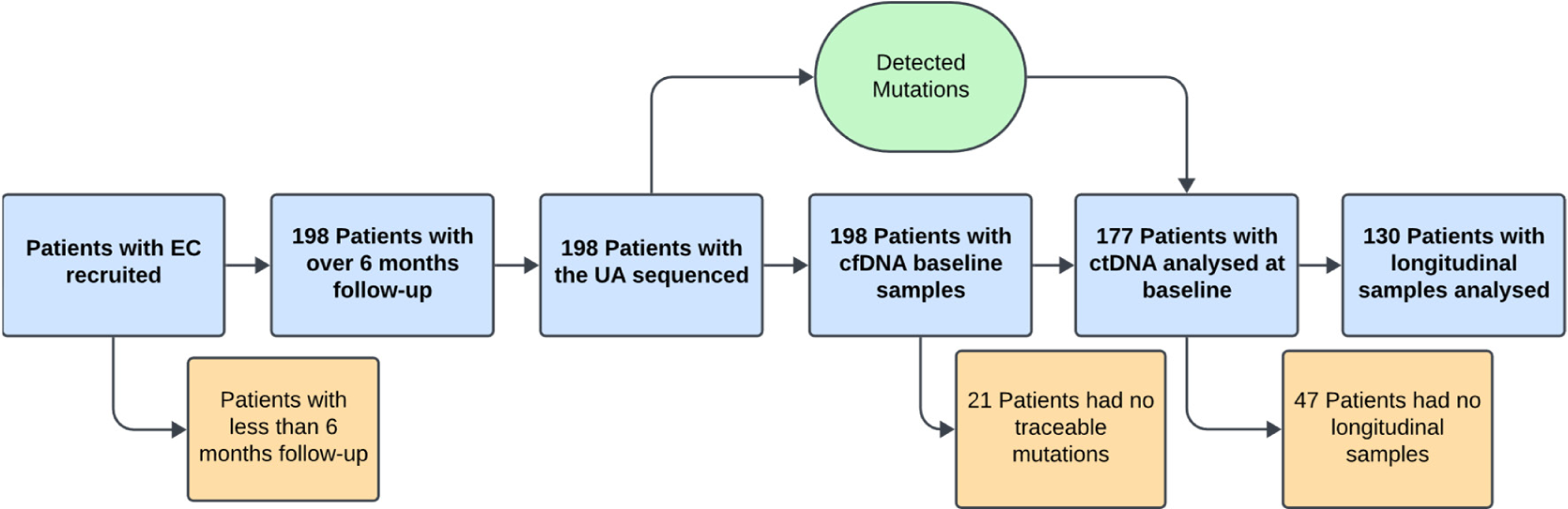
Design of the cfDNA and ctDNA analyses in the cohort of endometrial cancer patients. Consort diagram showing the number of patients excluded at each time point of the study.

### 3.3. Pre-surgery CfDNA levels have independent prognostic value in endometrial cancer patients

The total cfDNA concentration was evaluated to determine its association with the pathological findings and risk of recurrence. Total cfDNA was isolated from plasma samples (3-5mL) obtained at the time of tumour resection and quantified using Qubit fluorometry. The total cfDNA concentration at surgery ranged between 3.62-366.80 ng/mL with a mean value at 21.94 ng/mL and a median of 15.12 ng/mL. Higher levels of cfDNA correlated with traditional high risk of recurrence markers, although statistically significance was only found for myometrial and lympho vascular infiltration (Mann Whitney test p-value>0.05) (Figure 2A-H). Accordingly, significant higher pre-surgery cfDNA levels were found in patients who showed disease relapse or died because of disease (Mann Whitney test p-value<0.01) (Figure 2I-J). A cut-off at 25ng/mL was stablished to group patients into high or low cfDNA levels and explore the utility of cfDNA as a predictor of clinical outcome. Following this strategy 20.70% (41/198) of patients showed high pre-surgery cfDNA levels (Figure 2J). These patients had a significantly shorter DFS (Log-rank test p-value<0.0001; HR=3.91; 95% CI [2.04-7.51]) and DSS (Log-rank test p-value<0.0001; HR=6.54; 95% CI [2.83-15.10]) than those with low levels of pre-surgery cfDNA (Figure 2L-M, respectively). Moreover, multivariant analyses showed that cfDNA levels had independent prognostic value to predict DFS (Log-rank test p-value=0.008; HR=2.98; 95% CI [1.35-6.61]) and DSS (Log-rank test p-value<0.001; HR=9.13; 95% CI [2.82-29.50]) (Supplementary Table 3). Moreover, the correlation between cfDNA levels and standard blood biomarkers used for follow-up in the clinical setting, such as CA-125 or CEA, was analysed in part of the cohort. Importantly, cfDNA levels did not correlate with CA-125 and CEA levels (Spearman R<0.1) (Supplementary Figure 1A-C).

**Figure 2:**
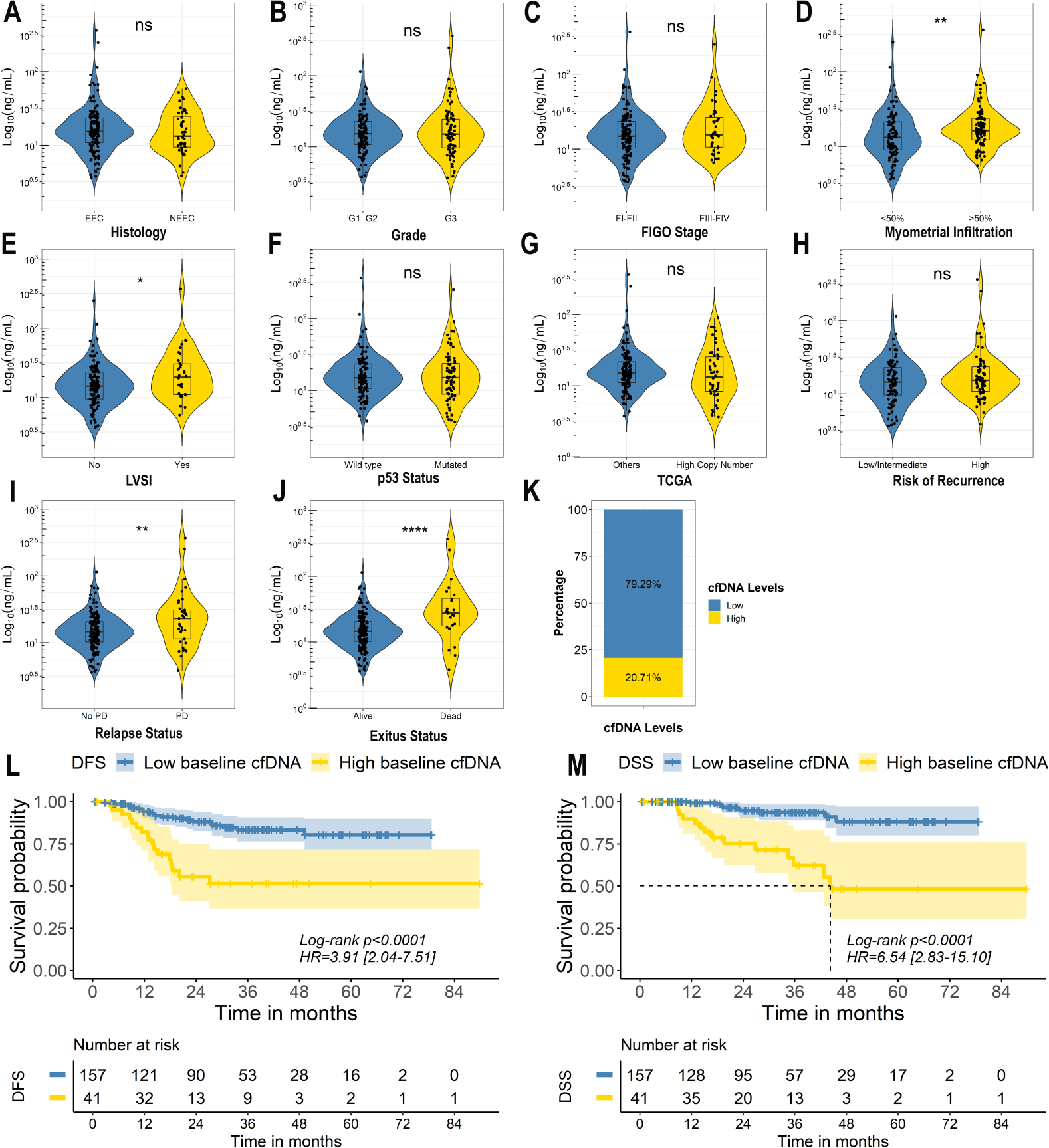
The value of pre-surgery cfDNA to identify patients with poor clinical outcome. **A-J** Violin plots of the pre-surgery cfDNA levels (Log_10_ ng/mL) according to the clinicopathologic variables of the tumours. Statistical significance was assessed based on Mann–Whitney U test **p<0.01. **K** Classification of the patients as low or high pre-surgery cfDNA based on the optimal cut point (25 ng/mL). **L-M.** Kaplan Meier curves showing DFS (**L**) and DSS (**M**) based on pre-surgery cfDNA levels. Univariate Cox proportional-hazards model was used to estimate HR and log-rank test to report p-value.

No correlation between cfDNA levels and leucocytes count was found, suggesting that DNA released by leucocytes did not have a main impact on the total cfDNA levels in our cohort as published for other tumour types (30). Furthermore, to ensure that the value of cfDNA and ctDNA were not biased towards tumour size or volume we performed a comparison of cfDNA levels and ctDNA positivity with tumour length (Supplementary Figure 1D-F, respectively) and with tumour volume (Supplementary Figure 1E-G, respectively), finding no differences between these variables.

### 3.3 CtDNA as a minimally invasive prognostic tool in endometrial cancer patients

The levels of ctDNA were determined in a total of 177 patients using personalized ddPCR assays based on the mutational profile found in the UA and 52 (29.38%) of them showed detectable levels of ctDNA (Figure 3K) with a variant allelic frequency (VAF) in a range from 0.01-39.10%, an average of 4.08% and a median of 0.44%. Pre-surgery ctDNA positivity was significantly associated with higher levels of cfDNA (Mann Whitney, p-value<0.01) (Supplementary Figure 2A) being this association partially explained by a lower cfDNA input used for the ddPCR in patients with low cfDNA (supplementary Figures 2B). However, pre-surgery cfDNA levels and ctDNA VAF were not correlated in those patients with detectable ctDNA levels (Spearman R≤0.2) (Supplementary Figure 2C-D). Higher detection rates and VAFs were detected in tumours with clinico pathological features of high risk, more specifically in patients with high grade, FIGO III-IV, over 50% myometrial infiltration or LVSI (Mann Whitney, p-value<0.01) (Figure 3A-H) (Supplementary Table 2), confirming that higher risk EC tumours shed into circulation higher ctDNA contents.

**Figure 3:**
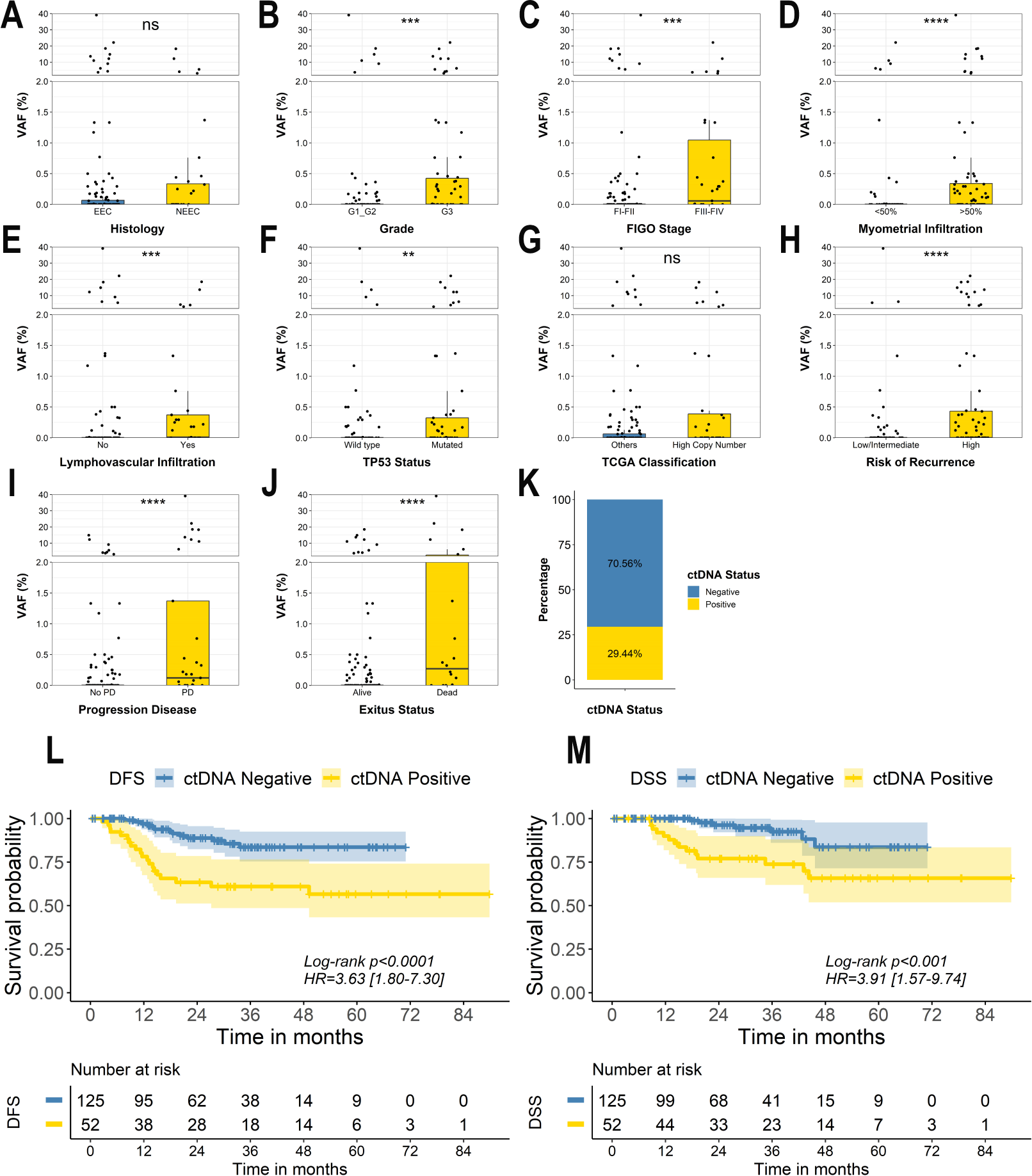
The value of ctDNA analyses in endometrial cancer. **A-J** Box plots representing the highest variant allelic frequency (VAF %) of the alterations found in the ctDNA accordingly the patient clinical variables. Statistical significance was assessed based on Mann–Whitney U test *p<0.05, **p<0.01, ****p<0.001. **K** Percentage of patients with positive and negative levels of ctDNA. **L-M.** Kaplan Meier curves showing DFS (**L**) and DSS (**M**) in patients with positive vs negative levels of ctDNA. Univariate Cox proportional-hazard model was used to estimate HR and log-rank test to report p-value.

With the aim to understand if pre-surgery ctDNA can provide additional information to predict the disease prognosis we grouped patients in positive and negative for ctDNA presence and performed survival analyses. Of note, patients with detectable levels of ctDNA showed significant shorter DFS (Log-rank test p-value<0.001; HR=3.63; 95% CI [1.80-7.30]) and DSS (Log-rank test p-value<0.01; HR=3.91; 95% CI [1.57-9.74]) when compared to patients with undetectable ctDNA (Figure 3L-M, respectively) (Supplementary Table 4).

### 3.4 Combinatory analysis of cfDNA and ctDNA identifies patients with worst clinical outcomes

Since cfDNA and ctDNA levels independently provided prognostic information, we aimed to explore if by combining both we could improve the identification of patients at higher risk of EC recurrence. With this purpose, patients were classified according to cfDNA levels and the presence/absence of ctDNA at surgery. Using this approach 4 groups were set up: ‘Group 1’ cfDNA-Low/ctDNA-Negative (58.76%); ‘Group 2’ cfDNA-Low/ctDNA-Positive (20.34%); ‘Group 3’ cfDNA-High/ctDNA-Negative (11.86%) and ‘Group 4’ cfDNA-High/ctDNA-Positive (9.04%). Clinical characteristics of the patients included in each group are summarised on Supplementary table 5.

Patients in group 4 (cfDNA-High/ctDNA-positive) had the worst results in terms of DFS (Log-rank test p-value<0.0001; HR=9.25; 95% CI [4.49-19.10]) and DSS (Log-rank test p-value<0.0001; HR=11.20; 95% CI [4.73-26.60]) when compared to the remaining groups (Figure 4A-B, respectively). Patients in group 3 (cfDNA-high/ctDNA-negative) showed poorer survival rates than patients in group 1 (cfDNA-low/ctDNA-negative) but similar with the group 2 (cfDNA-low/ctDNA positive) (Supplementary Figure 3A-B). These results could be associated with the presence of false ctDNA negative patients included in group 3 due to the limitations of the ddPCR approach, although cfDNA high levels indicate an aggressive disease.

**Figure 4.**
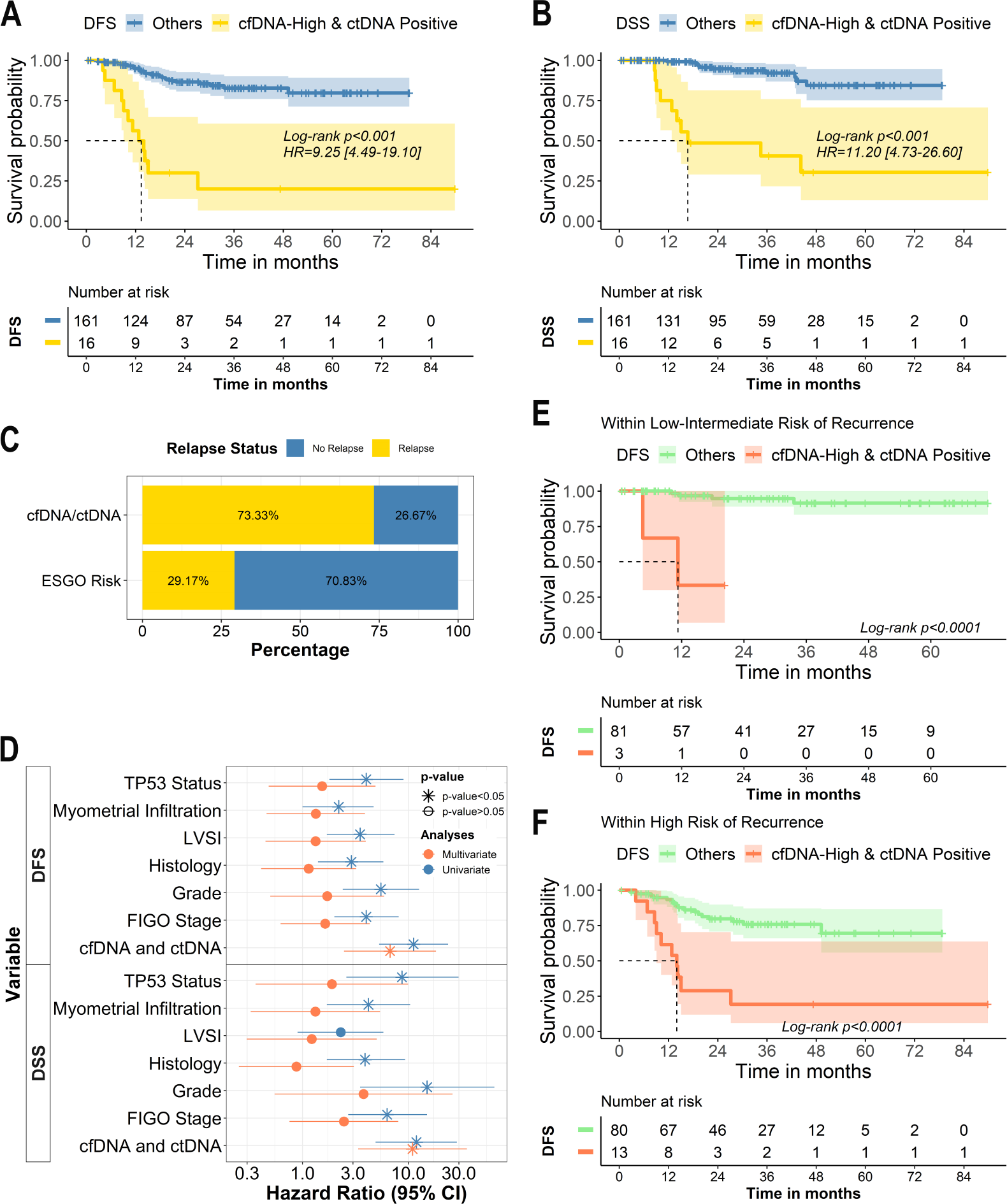
Combinatory analyses of cfDNA and ctDNA identify the patients with the worst clinical outcome. **A-B** Kaplan Meier curves showing DFS (**A**) and DSS (**B**) in patients according to the pre-surgery high levels of cfDNA and detectable levels of ctDNA. **C.** Bar plot with the early relapse status according to the combinatory approach and the ESGO risk classification **D.** Graphical representation of the univariate (blue) and multivariate (red) Cox proportional-hazard models. P-value>0.05 is represented with the *symbol. **E-F** Kaplan Meier curves showing DFS in patients according to the pre-surgery high levels of cfDNA and detectable levels of ctDNA in patients with low or intermediate risk (**E**) and high-intermediate or high risk (**F**) of recurrence based on the ESGO-ESTRO-ESP risk stratification.

Of note, 75% of patients included in group 4 (characterized by high cfDNA and ctDNA positivity) had a disease relapse, and 69% of them died during the follow-up as a result of the disease. Of note, 73% these patients showed a relapse within the first year after the surgery while only the 30% of the tumours classified as high-intermediate or high risk of recurrence, according to the latest ESGO-ESTRO-ESP risk classification, showed an early relapse (Figure 3C). Importantly, this combinatory approach showed independence over the traditional risk factors and molecular subtype (Figure 3D). Besides, this combinatory approach remains clinically significant when stratifying patients based on or according to histology, grade or FIGO stage (Supplementary Figure 3C-F). Of note, the presence of high levels of pre-surgery cfDNA and ctDNA positivity in patients classified as low or intermediate risk based on ESGO-ESTRO-ESP criteria was associated with a quick relapse in three cases, although most of the patients showed a good prognosis (Figure 4E). And also importantly, patients classified as high-intermediate/high risk of recurrence based on ESGO-ESTRO-ESP criteria and with high levels of pre-surgery cfDNA and the ctDNA positivity showed a very aggressive disease (Figure 4F). Therefore, the analysis of liquid biopsy clearly complements the current tools to anticipate disease relapse (Supplementary Figure 3G-H).

### 3.5. Combination of risk classification and cfDNA/ctDNA to predict the patients’ outcome

We combined the current risk stratification tools with the risk groups derived from the liquid biopsy analyses. With this strategy we considered a patient in the group of poor prognoses if she has a high-intermediate or high risk tumour (ESGO-ESTRO-ESP criteria) or high cfDNA/ctDNA positivity at surgery. This approach identified 54.23% (96/177) of the cohort as poor prognosis. With this approach the HRs of the poor prognosis group associated with the DFS (Log-rank test p-value<0.0001; HR=7.91; 95% CI [2.39-26.20]) and DSS (Log-rank test p-value<0.0001; HR=16.10; 95% CI [2.14-120]) were even more prominent than when analyse this classification strategies independently (Supplementary Figure 4A-B, respectively). Moreover, 90% and 95% of the patients who underwent disease recurrence and died because of the disease respectively, were classified as high risk patients thanks to the inclusion of the liquid biopsy in the analysis.

### 3.6 CfDNA and ctDNA as a monitoring tool for EC

To explore the value of cfDNA and ctDNA monitoring as a surrogate of the EC burden, a total of 372 longitudinal blood samples were analysed in a subset of 130 patients. From these patients, 22 showed disease progression. Significant reduction on the cfDNA levels were found after surgical resection at 1, 6 and 24 months (Wilcoxon signed-rank test, p-val<0.05) (Supplementary Figure 5A-F), although the dynamics of cfDNA levels in the longitudinal samples did not show value to anticipate the disease reappearance in our cohort of patients.

Notably, the analyses of the specific tumour fraction through longitudinal samples allowed for the identification of the disease recurrence months before (4.68±2.98) the clinical confirmation of relapse, mainly in patients with detectable pre-surgery ctDNA (80%, 8/10) (Figure 5A). However, in 4 (18.18%) of the 22 patients who showed recurrent disease, the ctDNA was not detectable with the ddPCR approach (Supplementary Figure 5G). Most of these patients were also ctDNA negative at surgery, showing the need to improve the sensitivity of the ctDNA detection to monitor tumours which are low shedders. Besides, only 3 of the 37 samples analyzed one month after surgery was positive. Accordingly in these patients the surgery was not radical, and presence of residual disease was known by the gynaecologists.

**Figure 5:**
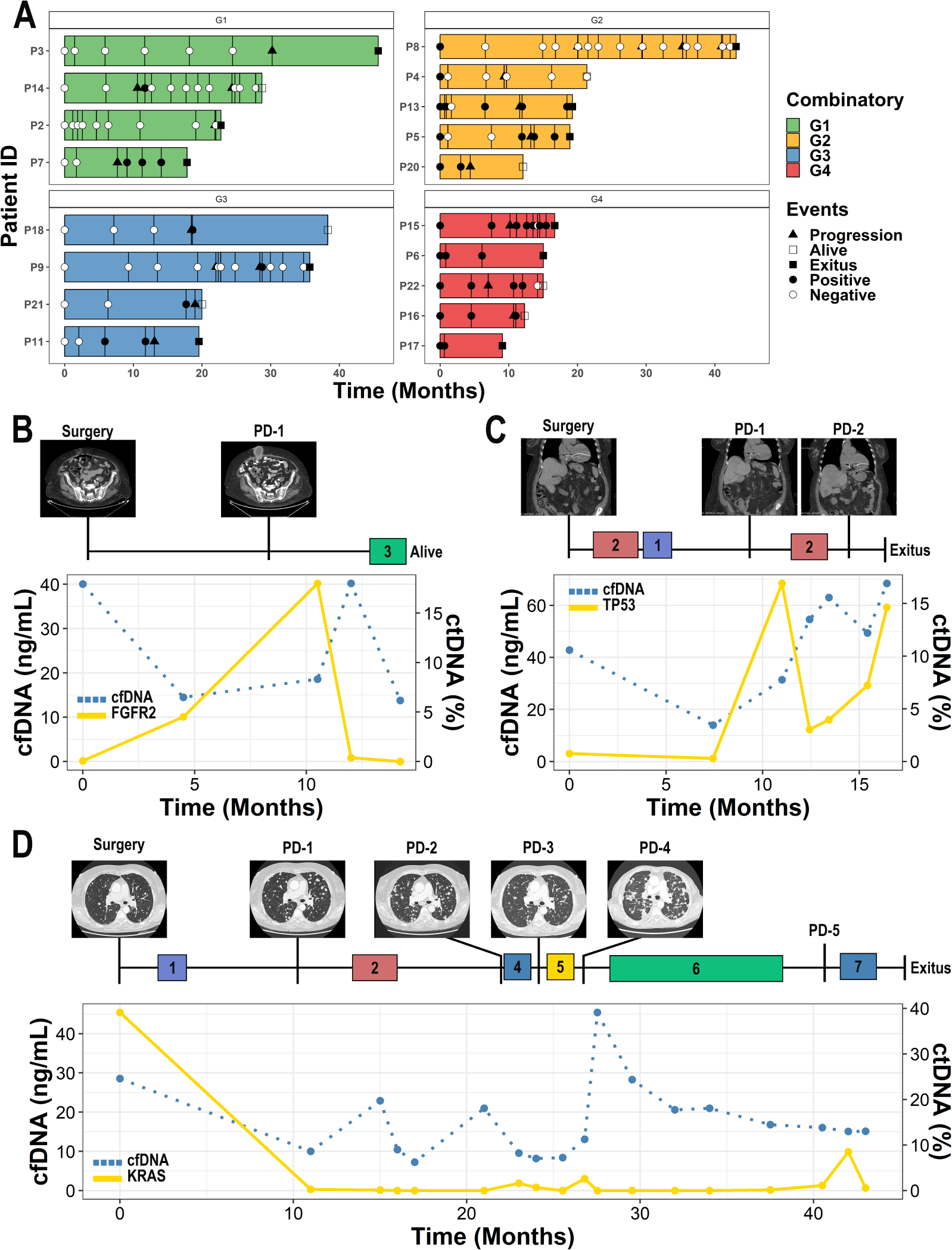
The longitudinal analyses of cfDNA and ctDNA allow for early detection of disease progression and accurately reflect the disease kinetics in response to treatment. **A.** Swimmer plot of the 18 patients that underwent tumour progression divided based on the combinatory approach with longitudinal samples collected at least 6 months prior to the relapse (cfDNA and ctDNA). **B-D.** Example figures of the cfDNA (blue dotted line) and ctDNA kinetics (yellow line) in patients with advanced disease. 1. Radiotherapy, 2. Carboplatin-Paclitaxel, 3. Dostarlimab, 4. Exemestane. 5. Doxorrubicin and Avastin. 6. Lenvatinib and Pembrolizumab, 7. Topotecan and Bevacizumab.

Importantly, longitudinal analyses of ctDNA proved to be a powerful asset to identify patients undergoing an early relapse, as reflected the patient #1 described in Figure 5B. This patient was diagnosed with an Endometrioid FIGO Stage IB tumour, positive for a pathogenic mutation in *FGFR2* in the pre-surgery cfDNA (0.06%), which was found increased 5 months after surgery (4.52%) and two months later a relapse at abdominal level was clinically confirmed (18%). The patient started to receive Dostarlimab and the ctDNA strongly decreased (0.38%) in line with the partial response defined based on CT-Scan. The patient is currently in response (0%) and being monitored by means of the ctDNA together with the image assessment.

Longitudinal analyses also allow for the dynamic characterization of the disease in response to therapy pressure (Figure 5C-D). For example, in Figure 5C is represented the case of a patient that was diagnosed with EC of serous histology of high grade and FIGO IIIA. NGS analyses of the UA showed alterations within the *PPP2R1A* and *TP53* genes and they were followed through the course of the disease. The patient had high levels of cfDNA at surgery as well as detectable levels of ctDNA. According to clinical guidelines the patient was treated with carboplatin-paclitaxel combined therapy and radiotherapy. Afterwards, the ctDNA levels were measured again, finding still detectable levels that were indicative of persistence of the disease (0.29%). Shortly after, the patient was confirmed to have a disease relapse at the liver through CT-Scan and showed a spike on ctDNA levels (16.90%). After the recurrence, patient was treated with a second-line chemotherapy. Although there was an initial reduction on ctDNA levels, they started to increase again and the patient showed progressive disease with peritoneal affectation and entered PS-ECOG 4 and could no longer be treated.

Another example is reflected on Figure 5D, a patient diagnosed with a mixed histology phenotype (initially diagnosed as endometrioid), grade 2, FIGO IB tumour that showed high levels of cfDNA and ctDNA at surgery. After a year, patient showed symptoms compatible with a disease relapse at the lungs. At this moment ctDNA was positive confirming the recurrence of the disease. Due to the nature of the relapse the patient was closely monitored throughout the course of the disease, and the cfDNA/ctDNA kinetics reflects the evolution of the disease and the response to therapy, being able to identify the resurge of the disease prior to clinical evidence. Thanks to this approach, clinicians have been able to adjust the treatment and anticipate CT-Scans according to the tumour kinetics. It is important to note that traditionally clinical variables identified this patient as being of intermediate risk, but our combinatory approach of cfDNA and ctDNA analyses classified it as of being of high risk of recurrence, reinforcing the additional value of liquid biopsy to anticipate the disease relapse.

### 4. DISCUSSION

To our knowledge, the present study represents the largest multicentre study conducted on EC that assesses the value of cfDNA and ctDNA as minimally invasive biomarkers to predict patient outcome at the moment of diagnosis but also to evaluate how longitudinal samples may help in the follow-up of EC patients. We have shown that cfDNA and ctDNA levels can help to identify patients with shorter DFS and DSS in pre-surgical setting and therefore tailor the adjuvant treatment. In addition, we have shown that close follow-up of ctDNA in patients at high risk of recurrence allows for the early detection of the disease relapse.

Although cfDNA origin is still unclear, multiple release pathways have been proposed, such as through apoptosis, necrosis, NETosis or from extracellular vesicles, among others (31,32). Most of the plasma cfDNA found in healthy people is thought to be derived by nucleated blood cells, such as neutrophils and lymphocytes (31). CfDNA levels are influenced by multiple factors such as age, metabolic activity, immune processes and disease, such as cancer (33). In patients with different solid tumour types cfDNA levels are found increased in comparison with healthy people and has been associated with poor prognosis in advanced stages (30,34) and, appart from the contribution of the tumour derived cfDNA in this increment, the neoplasic transformation may have systemic effect on cell turnover or DNA clearance associated with the cfDNA dynamic (30). In the present study we have shown that pre-surgery assessment of cfDNA levels correlates with poor clinical outcome, being a robust and independent prognostic biomarker for EC patients. We found a trend to have higher levels of cfDNA in high grade and myometrial/lymph vascular invasion as it was previously described by our group and other groups with different methodologies (21,26,35). These high cfDNA levels in high-risk tumours can be partially explained by a higher ctDNA release but also other systemic mechanisms which are more intensely regulated in advanced stages of the disease. In our study no correlation between cfDNA and the pre-surgery levels of different blood cells population was found, but we could not rule out the impact of this population on the cfDNA content without the application of more specific analyses to characterize the cfDNA origin in our EC population. Nevertheless, data presented clearly pointed to the cfDNA analysis as simple and cost-effective biomarker with prognostic value at surgery.

Indeed, when we specifically analyse the ctDNA fraction through personalized ddPCR assays to track MSI markers or pathogenic alterations (SNVs or CNVs) identified in the UAs, we found detectable levels in the 38% of the global cohort of patients, being this positivity more frequent in high grade (41.10%) or deep infiltrating tumours (46.91%). Regarding the molecular EC subtypes, *TP53* mutant tumours also showed higher detection of pre-surgery ctDNA. VAFs were found higher in tumours with high-risk characteristics. These data are in line with previous studies which demonstrated a high ctDNA content linked to high risk or advanced endometrial tumours (19,21,23,26,36). Accordingly, we found that patients with detectable levels of ctDNA at surgery had significantly shorter DFS and DSS times, although with no independent value over the rest of clinical variables, probably due to the strong correlation with the other risk factors under study.

The combination of both high levels of cfDNA and detectable ctDNA data at surgery allowed for the identification of a group of patients that showed an extremely poor clinical outcome. Although this group represents the 10% of the global cohort of patients analysed in our study, most of them developed an early relapse within the first year after surgery. The combined analysis of the pre-surgery cfDNA and ctDNA served to discriminate the risk of recurrence independently the histology, the FIGO stage, the grade or the molecular subtype. In fact, combined DNA analyses served to identify the patients that will recur within the groups of low, intermediate-high/ high risk tumours of ESGO/ESTRO/ESP classification. It is important to mention that patients with low levels of cfDNA and detectable levels of ctDNA at surgery showed a poor evolution in comparison with the patients with low cfDNA and undetectable ctDNA. However, this group was not so clinically relevant in comparison with the high cfDNA and positive ctDNA. We hypothesize that high cfDNA levels are indicating local or systemic changes associated with the tumour aggressiveness, non-directly linked to the ctDNA release, but impacting on the disease biology.

Our data demonstrate the clinical interest of cfDNA/ctDNA analysis at surgery to improve current risk stratification in patients with EC. In low and intermediate risk patients, although few cases were reclassified with the liquid biopsy approach, their DFS was significantly shorter. In addition, we consider also really relevant the ctDNA assessment in patients with high cfDNA or other risk factors, since the ctDNA presence will be a key tool to define the cohort of patients that would benefit from a closer follow-up or an intensification of the adjuvant treatment.

Notably, the ctDNA monitoring represents a valuable approach to detect the presence of minimal residual disease, anticipate relapse and evaluate the response to the therapy in advanced EC as the present study and other works have evidenced (21,27,37). From the total cohort of patients included in the study, we monitored the cfDNA and ctDNA levels in longitudinal plasma samples from 130 patients. Globally, the cfDNA dynamic lacked value to accurately mirror the tumour burden although in specific cases cfDNA levels changed accordingly with the tumour evolution. This result could be explained by the addition of other factors such as the adjuvant therapy that can modify systemically the cfDNA release into circulation (38). Of note, in the follow-up setting ctDNA monitoring showed high value to track the disease evolution as evidence the particular cases described. Post-surgery ctDNA was only detected in cases with residual or recurrent disease. Specially in cases with presence of pre-operative ctDNA, the ctDNA kinetics served to detect the relapse months before the clinical/radiological confirmation, providing an opportunity to start the treatment earlier and accounting with the molecular information of the tumour clones that are driving the recurrent disease. The clinical relevance of post-surgery ctDNA monitoring has been well documented in other tumour types like colorectal, lung or breast tumours (39–42) and also in EC longitudinal ctDNA assessment by NGS and ddPCR has been successfully applied to detect the disease progression (21,27,37).

Our study represents the larger cohort of EC patients monitored with ctDNA so far, and robustly demonstrates the benefit of this strategy to make a more personalized and precise disease follow-up. Although these positive results, our approach has some limitations since ctDNA pre/post-surgery detection was not efficient in 20% patients who showed disease relapse. To this regards it’s important to mention that our approach prioritise sensitivity by analysing the most predominant alterations found in the primary tumour of each patient; nonetheless, it may not be the predominant clone being released into the bloodstream and the arisen of new mutations within tumour evolution is not taken into consideration. Other technologies, such as panel-based strategies circumvent this limitation at the cost of sensitivity (21,37). Another study’s weakness is the lack of knowledge about the origin of the cfDNA found within the different sampling points. No correlation was found between blood cells levels and the cfDNA dynamics, but this cannot discard their contribution without other type of molecular characterization.

Although these limitations, the present study demonstrates the value of cfDNA and ctDNA analyses as prognostic tools in the largest EC cohort published so far. High levels of cfDNA and detectable levels of ctDNA at surgery strongly correlated with poor prognosis and served to identify the patients that suffered an early relapse independently of other known EC risk factors. Additionally, the longitudinal ctDNA assessment allowed for the early identification of recurrences and the development of resistance to the treatment. Implementation of this approach into the clinic would translate into a much better management of EC patients, reducing overtreatment and identifying patients at higher risk of recurrence for close monitoring. Although still far, our data suggest that the implementation of liquid biopsy into the clinic could greatly improve the management of the EC patients.

## AUTHOR CONTRIBUTIONS

L.M-R, M.A, A.G-M, G.M-B. and E.C. conceived and designed the study. C.C-A, A.A, E.D, C.P.M, C.L.G, S.C, E.A, V.S, J.C, A.C, T.C, M.P-S, S. DdP, P. P-I, M.A, A.H., V.G-P contributed to the collection of samples and clinical data. E.D, S.S.O, and G.M-B. performed the NGS analyses. C.C-A. carried out the cfDNA/ctDNA and statistical analyses. A.G-M, J.R-B, X.M-G and R.L contributed with the clinical interpretation of the experimental findings. C.C-A. and L.M-R. wrote the manuscript. All authors provided critical feedback and helped guide the research, analysis, and manuscript. All authors have read and agreed to the published version of the manuscript.

## ETHICS STATEMENT

The study was approved by the corresponding Research Ethics Committees (Galician Research Ethics Committee—reference number 2017/530 and 2022/029, Vall d’Hebron Research Ethics Committee—reference number PRAMI276-2018) and conducted in accordance with the guidelines for Good Clinical Practice and the Declaration of Helsinki. All patients provided written informed consent before enrolment.

## DATA AVAILABILITY STATEMENT

The datasets used and/or analyzed during the current study are avail-able from the corresponding author on reasonable request.

## ABBREVIATIONS

cfDNA: circulating free DNA
ctDNA: circulating tumour DNA
EC: Endometrial Cancer
HCN: High Copy number
HR: Hazards Ratio
LVSI: Lymphovascular space invasion
MSI: Microsatellite Instability
NSMP: Non Specific Molecular Profile
DSS: Disease Specific Survival
DFS: Disease Free Survival
ROC: Receiver operator curve
TCGA: The Cancer genome Atlas
UA: Uterine aspirate
VAF: Variant Allelic frequency

