## Supplementary for "ROLE OF cfDNA AND ctDNA TO IMPROVE THE RISK STRATIFICATION AND THE DISEASE FOLLOW-UP IN PATIENTS WITH ENDOMETRIAL CANCER: TOWARDS THE CLINICAL APPLICATION"

#### SUPLEMENTARY INFORMATION

**Supplementary Table 1.** Clinical characteristics of the studied cohort.

| CLINICAL VARIABLES | *N=198* | CLINICAL VARIABLES | *N=198* |
|---|---|---|---|
| **Age** | 181 | **Lymphovascular Infiltration** | 171 |
| Median (IQR) | 67 (58-73) | No | 137 (80%) |
| **Histology** | 196 | Yes | 34 (20%) |
| NEEC | 48 (24%) | **TCGA** | 194 |
| EEC | 148 (76%) | POLE | 15 (7.7%) |
| **Tumour Grade** | 197 | MSI | 76 (27%) |
| Grade 1 | 78 (40%) | NSMP | 53 (27%) |
| Grade 2 | 36 (18%) | HCN | 50 (26%) |
| Grade 3 | 83 (42%) | **cfDNA Concentration (ng/mL)** | 198 |
| **FIGO Stage** | 193 | Median (IQR) | 15 (10,24) |
| I | 126 (65%) | **ctDNA Positivity** | 177 |
| II | 29 (15%) | Detected | 52 (29%) |
| III | 29 (15%) | **Progression Disease** | 198 |
| IV | 9 (4.7%) | No | 161 (81%) |
| **Myometrial Infiltration** | 195 | Yes | 37 (19%) |
| <50% | 103 (53%) | **Death of Disease** | 198 |
| >50% | 92 (47%) | No | 174 (88%) |
| | | Yes | 24 (12%) |

**Supplementary Table 2.** Correlation between the presurgical cfDNA levels and ctDNA detection and different clinicopathologic features.

| **Variable** | **cfDNA (ng/mL)[1]** | **p-value** | **ctDNA Positivity** | **ctDNA VAF (%)[1]** | **p-value** |
|---|---|---|---|---|---|
| **Histology** | | | | | |
| EEC | 15 (11-23) | 0.5[2] | 28.15% (38/136) | 0 (0-0.07) | 0.3[2] |
| NEEC | 13 (9-25) | | 35.00% (14/40) | 0 (0-0.34) | |
| **Grade** | | | | | |
| G1-G2 | 15 (11-23) | >0.9[2] | 21.36% (22/103) | 0 (0-0) | **0.002**[2] |
| G3 | 15 (10-24) | | 41.10% (30/73) | 0 (0-0.44) | |
| **FIGO** | | | | | |
| I-II | 15 (10-23) | 0.3[2] | 23.18% (32/138) | 0 (0-0) | **<0.001**[2] |
| III-IV | 16 (11-26) | | 52.95% (18/34) | 0.04 (0-1.19) | |
| **Myometrial infiltration** | | | | | |
| <50% | 13 (9-21) | **0.005**[2] | 13.98% (13/93) | 0 (0-0) | **<0.001**[2] |
| >50% | 16 (12-24) | | 46.92% (38/86) | 0 (0-0.37) | |
| **LVSI** | | | | | |
| No | 15 (10-23) | **0.025**[2] | 18.85% (23/122) | 0 (0-0) | **<0.001**[2] |
| Yes | 20 (11,31) | | 46.67% (16/30) | 0.07 (0-0.42) | |
| **TCGA** | | | | | |
| POLE | 15 (11-18) | 0.6[3] | 17.67% (3/14) | 0 (0-0) | 0.10[3] |
| MSI | 15 (12-23) | | 35.13% (26/74) | 0 (0-0.16) | |
| NSMP | 15 (11-21) | | 17.78% (8/45) | 0 (0-0) | |
| HCN | 13 (8-25) | | 37.50% (15/40) | 0 (0-0.39) | |
| **Risk of recurrence** | | | | | |
| Low/intermediate low | 14 (10-23) | 0.2[2] | 16.67% (14/84) | 0 (0-0) | **<0.001**[2] |
| High/high-intermediate | 15 (11-23) | | 40.86% (55/93) | 0 (0-0.4) | |
| **Relapse** | | | | | |
| No | 15 (10-21) | **0.008**[2] | 22.22% (32/144) | 0(0-0) | **<0.001**[2] |
| Yes | 23 (11,31) | | 60.61% (20/33) | 0.12 (0-1.37) | |
| **Death of Disease** | | | | | |
| No | 15 (10-21) | **<0.001**[2] | 24.36% (38/156) | 0 (0-0) | **<0.001**[2] |
| Yes | 27 (17,44) | | 66.67% (14/21) | 0.22 (0-1.37) | |

[1]Median (IQR)
[2]Wilcoxon rank sum test
[3]Kruskal-Wallis rank sum test

**Supplementary Table 3. cfDNA analyses identify the patients with the worst clinical outcome.** Cox proportional-hazards model was used to determine the relationship between clinical variables and the experimental variables.

| | Univariate | | | | | Multivariate | | | |
|---|---|---|---|---|---|---|---|---|---|
| **Variable** | **N** | **HR**[1] | **95% CI**[1] | **p-value** | **q-value**[2] | **HR**[1] | **95% CI**[1] | **p-value** | **q-value**[2] |
| **Disease Free Survival** | | | | | | | | | |
| Histology | 196 | 3.72 | 1.95,7.11 | **<0.001** | **<0.001** | 2.16 | 0.83, 5.65 | 0.10 | 0.37 |
| Grade | 197 | 5.42 | 247,11.9 | **<0.001** | **<0.001** | 1.84 | 0.58, 5.81 | 0.30 | 0.37 |
| FIGO Stage | 193 | 4.04 | 2.10, 7.79 | **<0.001** | **<0.001** | 1.68 | 0.68, 4.17 | 0.26 | 0.37 |
| Myometrial Infiltration | 195 | 2.36 | 1.16, 4.77 | **0.013** | **0.015** | 1.31 | 0.49, 3.51 | 0.60 | 0.60 |
| LVSI | 171 | 4.26 | 2.17, 8,38 | **<0.001** | **<0.001** | 1.72 | 0.65, 4.55 | 0.28 | 0.37 |
| MSI Status | 175 | 0.62 | 0.30, 1.27 | 0.21 | 0.21 | | | | |
| TP53 Status | 188 | 4.08 | 1.90, 8.75 | **<0.001** | **<0.001** | 1.66 | 0.60, 4.57 | 0.32 | 0.37 |
| cfDNA Levels | 198 | 3.91 | 2.04, 7.51 | **<0.001** | **<0.001** | 2.98 | 1.35, 6.61 | **0.008** | **0.058** |
| **Disease Specific Survival** | | | | | | | | | |
| Histology | 196 | 4.46 | 1.95,10.2 | **<0.001** | **<0.001** | 1.05 | 0.34,3.32 | 0.93 | 0.93 |
| Grade | 197 | 16.4 | 3.84,70.2 | **<0.001** | **<0.001** | 4.75 | 0.85,26.5 | 0.056 | 0.13 |
| FIGO Stage | 193 | 7.10 | 3.07,16.4 | **<0.001** | **<0.001** | 4.35 | 1.30,14.6 | **0.015** | **0.054** |
| Myometrial Infiltration | 195 | 2.54 | 1.00,16.4 | **0.038** | **0.043** | 1.28 | 0.31,5.35 | 0.73 | 0.93 |
| LVSI | 171 | 4.90 | 2.07,11.6 | **<0.001** | **<0.001** | 0.86 | 0.23,3.25 | 0.83 | 0.93 |
| MSI Status | 175 | 0.52 | 0.20,1.35 | 0.16 | 0.16 | | | | |
| TP53 Status | 188 | 9.32 | 2.74,31.6 | **<0.001** | **<0.001** | 3.05 | 0.72,12.9 | 0.11 | 0.19 |
| cfDNA Levels | 198 | 6.54 | 2.83,15.1 | **<0.001** | **<0.001** | 9.13 | 2.82,29.5 | **0.001** | **0.001** |

[1] HR = Hazard Ratio, CI = Confidence Interval

[2] False discovery rate correction for multiple testing

**Supplementary Table 4. ctDNA analyses allow for the identification of the patients with the worst clinical outcome.** A cox proportional-hazards model was used to determine the relationship between clinical variables and the experimental variables.

| | Univariate | | | | | Multivariate | | | |
|---|---|---|---|---|---|---|---|---|---|
| **Variable** | **N** | **HR[1]** | **95% CI[1]** | **p-value** | **q-value[2]** | **HR[1]** | **95% CI[1]** | **p-value** | **q-value[2]** |
| **Disease Free Survival** | | | | | | | | | |
| Histology | 196 | 3.72 | 1.95,7.11 | **<0.001** | **<0.001** | 2.57 | 0.99,6.69 | **0.047** | 0.16 |
| Grade | 197 | 5.42 | 247,11.9 | **<0.001** | **<0.001** | 1.26 | 0.37,4.25 | 0.71 | 0.75 |
| FIGO Stage | 193 | 4.04 | 2.10, 7.79 | **<0.001** | **<0.001** | 1.25 | 0.44,3.51 | 0.68 | 0.75 |
| Myometrial Infiltration | 195 | 2.36 | 1.16, 4.77 | **0.013** | **0.015** | 0.85 | 0.30,2.35 | 0.75 | 0.75 |
| LVSI | 171 | 4.26 | 2.17, 8,38 | **<0.001** | **<0.001** | 2.38 | 0.83,6.79 | 0.10 | 0.24 |
| MSI Status | 175 | 0.62 | 0.30, 1.27 | 0.21 | 0.21 | | | | |
| TP53 Status | 188 | 4.08 | 1.90, 8.75 | **<0.001** | **<0.001** | 1.33 | 0.45,3.99 | 0.61 | 0.75 |
| ctDNA Levels | 177 | 3.63 | 1.80,7.30 | **<0.001** | **<0.001** | 2.70 | 1.12,6.46 | **0.025** | 0.16 |
| **Disease Specific Survival** | | | | | | | | | |
| Histology | 196 | 4.46 | 1.95,10.2 | **<0.001** | **<0.001** | 2.11 | 0.71,6.31 | 0.18 | 0.35 |
| Grade | 197 | 16.4 | 3.84,70.2 | **<0.001** | **<0.001** | 2.56 | 0.39,16.9 | 0.31 | 0.39 |
| FIGO Stage | 193 | 7.10 | 3.07,16.4 | **<0.001** | **<0.001** | 2.25 | 0.66,7.65 | 0.19 | 0.35 |
| Myometrial Infiltration | 195 | 2.54 | 1.00,16.4 | **0.038** | **0.043** | 0.55 | 0.14,2.15 | 0.39 | 0.39 |
| LVSI | 171 | 4.90 | 2.07,11.6 | **<0.001** | **<0.001** | 2.32 | 0.62,8.72 | 0.20 | 0.35 |
| MSI Status | 175 | 0.52 | 0.20,1.35 | 0.16 | 0.16 | | | | |
| TP53 Status | 188 | 9.32 | 2.74,31.6 | **<0.001** | **<0.001** | 2.07 | 0.45,9.58 | 0.33 | 0.39 |
| ctDNA Levels | 177 | 3.91 | 1.57,9.74 | **0.002** | **0.003** | 3.40 | 1.18,9.77 | **0.018** | 0.13 |

[1] HR = Hazard Ratio, CI = Confidence Interval

[2] False discovery rate correction for multiple testing

**Supplementary Table 5.** Clinical characteristics of the cohort when divided based on the different combination groups.

| Variable | N | Group 1<br>N=104 | Group 2<br>N=36 | Group 3<br>N=21 | Group 4<br>N=16 | p-value[2] |
|---|---|---|---|---|---|---|
| **Age** | 160 | | | | | 0.60 |
| Median [IQR] | | 66 [56,73] | 69 [57,73] | 67 [62,75] | 69 [59,78] | |
| **Histology** | 175 | | | | | 0.20 |
| Endometrioid | | 79 (77%) | 30 (83%) | 18 (86%) | 8 (50%) | |
| Serous | | 16 (16%) | 4 (11%) | 2 (9.5%) | 8 (50%) | |
| Clear cell | | 3 (2.9%) | 0 (0%) | 0 (0%) | 0 (0%) | |
| Mixed | | 3 (2.9%) | 1 (3.0%) | 1 (4.8%) | 0 (0%) | |
| Carcinosarcoma | | 1 (1%) | 1 (2.8%) | 0 (0%) | 0 (0%) | |
| **Tumour Grade** | 176 | | | | | **<0.001** |
| 1 | | 53 (51%) | 7 (19%) | 9 (43%) | 0 (0%) | |
| 2 | | 13 (13%) | 11 (31%) | 6 (29%) | 4 (25%) | |
| 3 | | 37 (36%) | 18 (50%) | 6 (29%) | 12 (75%) | |
| **FIGO Stage** | 172 | | | | | |
| I | | 76 (75%) | 15 (44%) | 17 (81%) | 5 (31%) | |
| II | | 11 (11%) | 10 (29%) | 3 (14%) | 2 (13%) | |
| III | | 12 (12%) | 7 (21%) | 1 (4.8%) | 6 (38%) | |
| IV | | 2 (2%) | 2 (5.9%) | 0 (0%) | 3 (19%) | |
| **LVSI** | 152 | | | | | **<0.001** |
| No | | | | | | |
| Yes | | 9 (9.7%) | 10 (38%) | 5 (25%) | 6 (46%) | |
| **Myometrial Infiltration** | 174 | | | | | **<0.001** |
| <50% | | 69 (67%) | 7 (20%) | 11 (55%) | 6 (38%) | |
| >50% | | 34 (33%) | 28 (80%) | 9 (45%) | 10 (63%) | |
| **TCGA Classification** | 173 | | | | | |
| POLE | | 9 (8.80%) | 3 (8.30%) | 2 (11%) | 0 (0%) | |
| MSI | | 41 (40%) | 19 (53%) | 7 (37%) | 7 (44%) | |
| NSMP | | 31 (30%) | 7 (19%) | 6 (32%) | 1 (6.3%) | |
| HCN | | 21 (21%) | 7 (19%) | 4 (21%) | 8 (50%) | |
| **cfDNA Concentration** | 177 | | | | | **<0.001** |
| Median [IQR] | | 13 [9,16] | 15 [12,19] | 40 [28,56] | 31 [28,45] | |
| **ctDNA Positivity** | 177 | | | | | **<0.001** |
| | | 0 (0%) | 36 (100%) | 0 (0%) | 16 (100%) | |
| **Progression Disease** | 177 | | | | | **<0.001** |
| No | | 95 (91%) | 28 (78%) | 17 (81%) | 4 (25%) | |
| Yes | | 9 (8.7%) | 8 (22%) | 4 (19%) | 12 (75%) | |
| **Death of Disease** | 177 | | | | | **<0.001** |
| Alive | | 100 (96%) | 32 (89%) | 18 (86%) | 5 (31%) | |
| Dead | | 4 (3.8%) | 4 (11%) | 3 (14%) | 11 (69%) | |

[1]n (%)

[2]Kruskal-Wallis rank sum test; Fisher's exact test; Pearson's Chi-squared test

### SUPPLEMENTARY FIGURES

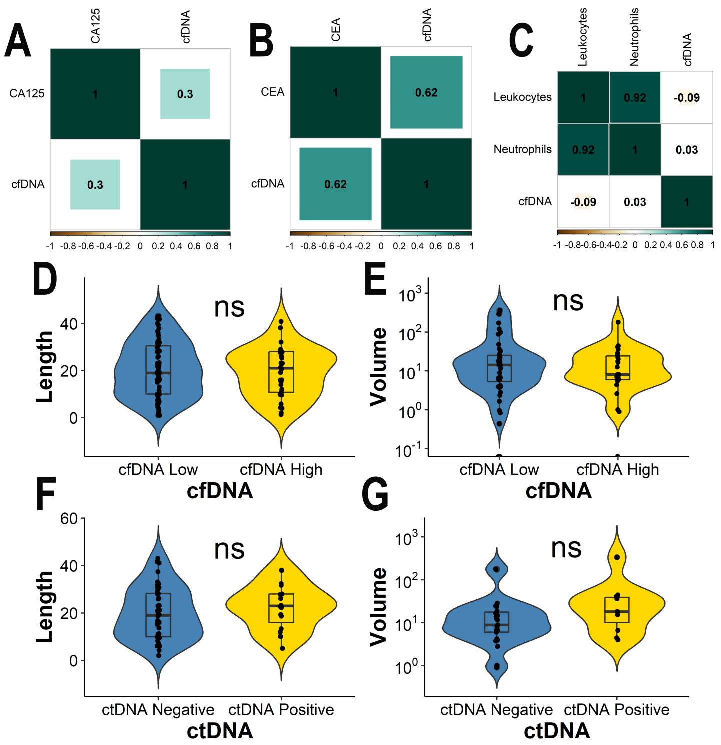


**Supplementary Figure 1:** Pre-surgery cfDNA and ctDNA levels showed independence of other blood parameters. **A-C** Spearman Correlation of the cfDNA levels with CA-125 (n=36) (**A**), CEA (n=18) (**B**) or PBMCs (n=58) levels (**C**). **D-G** Violin plots representing the length of the tumour (cm) according to the cfDNA levels (**D**) and ctDNA presence (**F**) and the tumour volume ($cm^2$) according the cfDNA levels (**E**) and ctDNA presence (**G**). Statistical significance was assessed based on Mann–Whitney U. ns p-value>0.05.

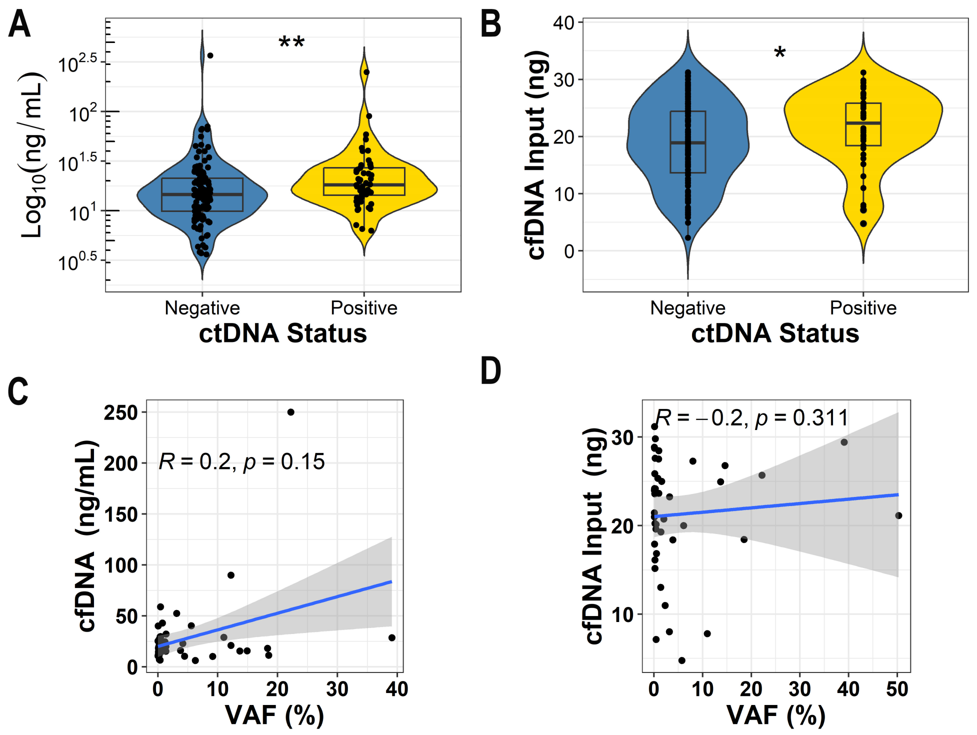


**Supplementary Figure 2: A** Violin plot of the cfDNA concentration according to the presence of ctDNA. **B** Violin plot of the cfDNA input used in the ddPCR according to the presence of ctDNA. **C** Spearman correlation between the cfDNA levels at baseline (ng/mL) and the ctDNA VAFs (%). **D** Spearman correlation of ctDNA levels (VAF=%) and cfDNA input used in the ddPCR.

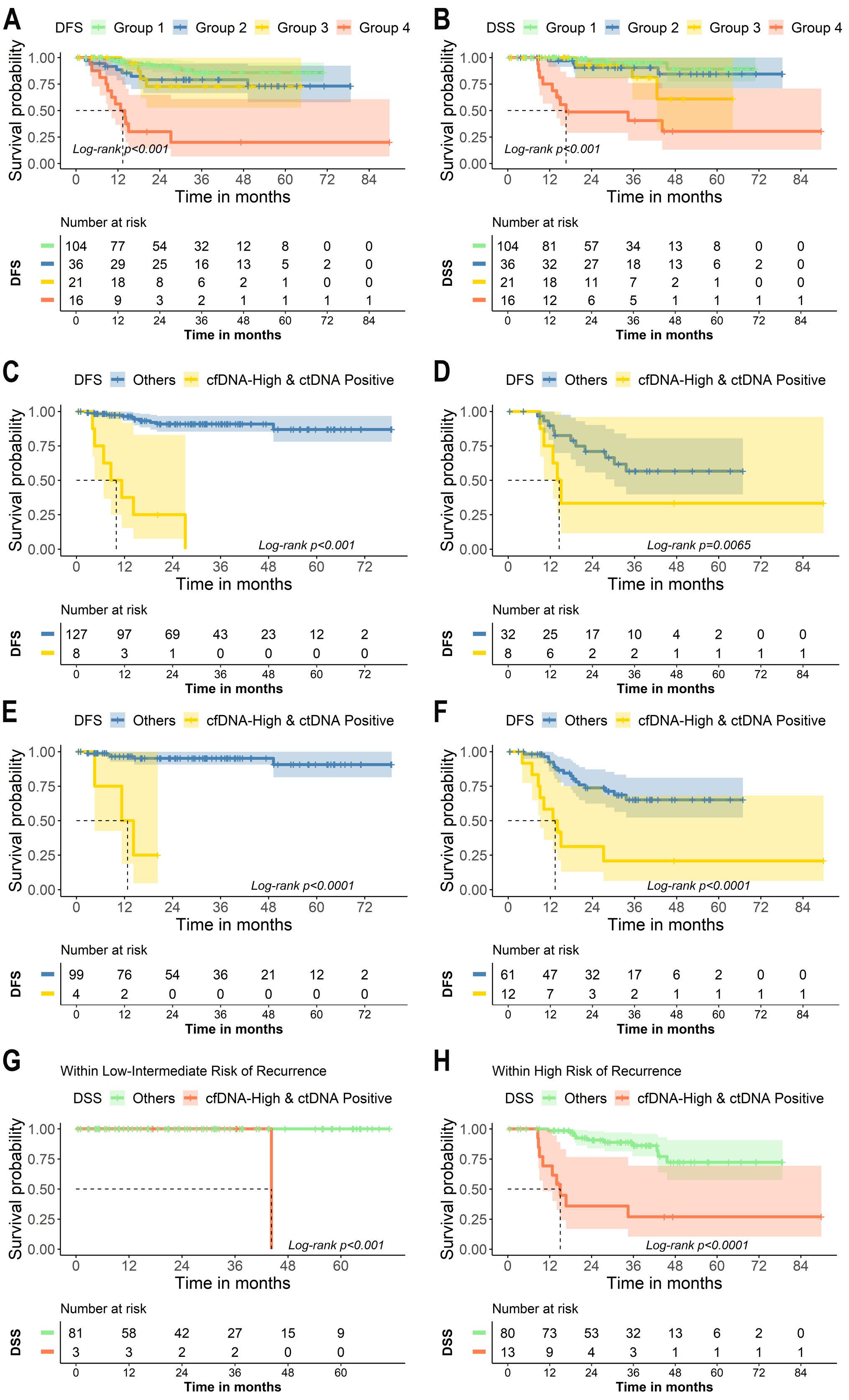
A
DFS Group 1 Group 2 Group 3 Group 4
Survival probability
Log-rank p<0.001
Time in months
Number at risk
DFS
104 77 54 32 12 8 0 0
36 29 25 16 13 5 2 0
21 18 8 6 2 1 0 0
16 9 3 2 1 1 1 1
0 12 24 36 48 60 72 84
Time in months
B
DSS Group 1 Group 2 Group 3 Group 4
Survival probability
Log-rank p<0.001
Time in months
Number at risk
DSS
104 81 57 34 13 8 0 0
36 32 27 18 13 6 2 0
21 18 11 7 2 1 0 0
16 12 6 5 1 1 1 1
0 12 24 36 48 60 72 84
Time in months
C
DFS Others cfDNA-High & ctDNA Positive
Survival probability
Log-rank p<0.001
Time in months
Number at risk
DFS
127 97 69 43 23 12 2
8 3 1 0 0 0 0
0 12 24 36 48 60 72
Time in months
D
DFS Others cfDNA-High & ctDNA Positive
Survival probability
Log-rank p=0.0065
Time in months
Number at risk
DFS
32 25 17 10 4 2 0 0
8 6 2 2 1 1 1 1
0 12 24 36 48 60 72 84
Time in months
E
DFS Others cfDNA-High & ctDNA Positive
Survival probability
Log-rank p<0.0001
Time in months
Number at risk
DFS
99 76 54 36 21 12 2
4 2 0 0 0 0 0
0 12 24 36 48 60 72
Time in months
F
DFS Others cfDNA-High & ctDNA Positive
Survival probability
Log-rank p<0.0001
Time in months
Number at risk
DFS
61 47 32 17 6 2 0 0
12 7 3 2 1 1 1 1
0 12 24 36 48 60 72 84
Time in months
G
Within Low-Intermediate Risk of Recurrence
DSS Others cfDNA-High & ctDNA Positive
Survival probability
Log-rank p<0.001
Time in months
Number at risk
DSS
81 58 42 27 15 9
3 3 2 2 0 0
0 12 24 36 48 60
Time in months
H
Within High Risk of Recurrence
DSS Others cfDNA-High & ctDNA Positive
Survival probability
Log-rank p<0.0001
Time in months
Number at risk
DSS
80 73 53 32 13 6 2 0
13 9 4 3 1 1 1 1
0 12 24 36 48 60 72 84
Time in months

**Supplementary Figure 3: A-B** Kaplan Meier curves showing DFS (**A**) and DSS (**B**) in patients according to the pre-surgery levels of cfDNA and ctDNA. Group 1: low presurgical levels of cfDNA and negative ctDNA, Group 2: low presurgical levels of cfDNA and positive ctDNA levels, Group 3: high presurgical levels of cfDNA and negative ctDNA, Group 4: high presurgical levels of cfDNA and detectable ctDNA levels. **C-F** Kaplan Meier curves showing DFS in patients according to the pre-surgery high levels of cfDNA and detectable levels of ctDNA in patients with EEC histology (**C**) NEEC histology (**D**) low grade (G1 or G2) (**E**) or high grade (G3) (**F**). **G-H** Kaplan Meier curves showing DSS in patients according to the pre-surgery high levels of cfDNA and detectable levels of ctDNA in patients with low/intermediate (**G**) or high-intermediate/high risk (**H**) of recurrence according to the ESGO stratification.

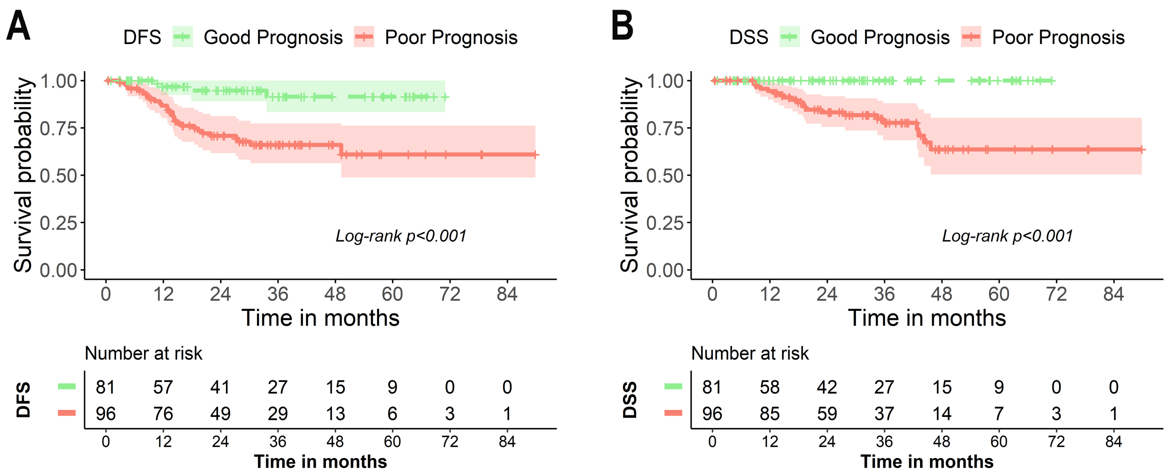


**Supplementary Figure 4: A-B** Kaplan Meier curves showing DFS (**A**) and DSS (**B**) in patients according to the pre-surgery levels of cfDNA and ctDNA and the risk of recurrence according to the ESGO stratification. Good prognosis is defined as patients of low or intermediate risk of recurrence according to the ESGO stratification and negative levels of ctDNA. Poor prognosis is defined as patients with either high-intermediate/high risk of recurrence according to the ESGO stratification or high levels of cfDNA and detectable levels of ctDNA regardless of the risk of recurrence.

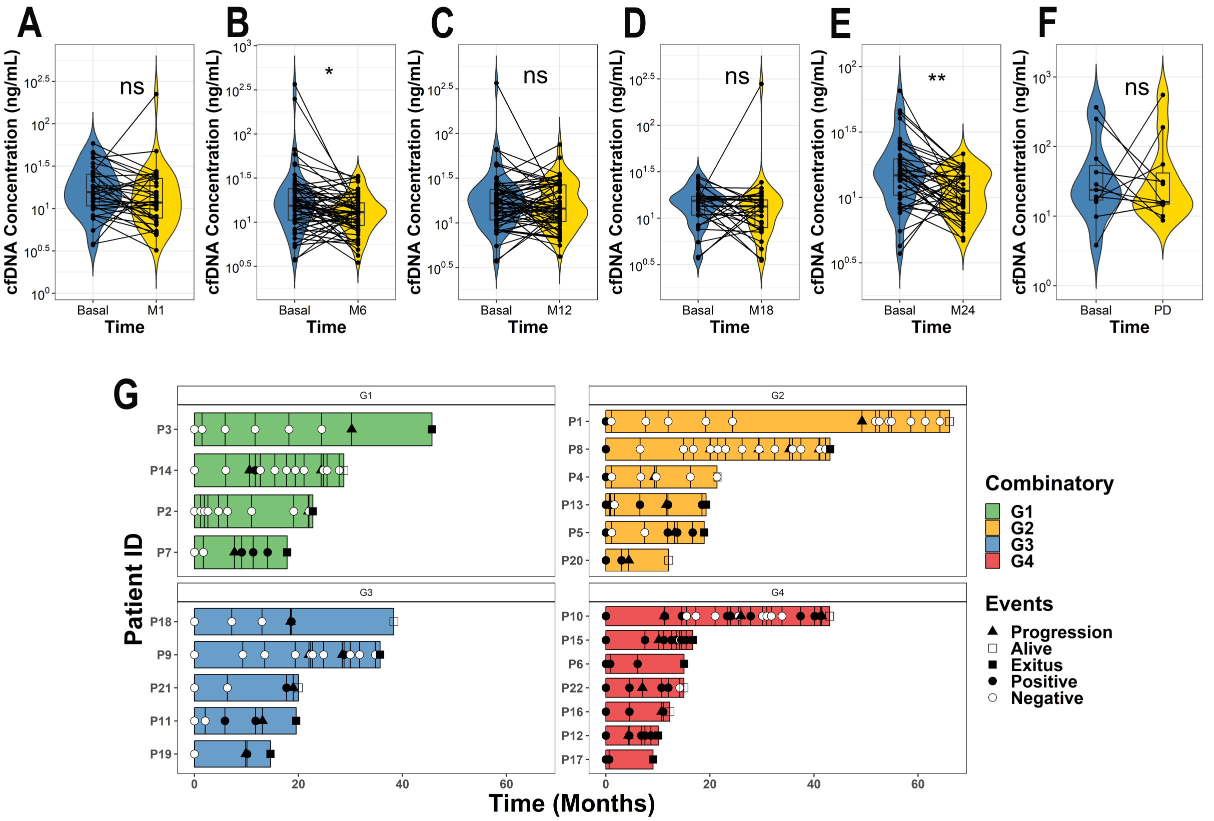


**Supplementary Figure 5: A-F** Violin plots of the cfDNA levels within the entire population (**A-F**) when compared between baseline and different time points: (**A**) 1 month post-surgery, (**B**) 6 months post-surgery, (**C**) 12 months post-surgery, (**D**) 18 months post-surgery, (**E**) 24 months post-surgery and (**F**) at progression disease. **G** Swimmer plot of all patients that underwent tumour progression divided based on the combinatory approach. Wilcoxon signed-rank test was used to assess statistical significance between paired samples *p<0.05.
